# The Association of Alzheimer’s Disease-related Blood-based Biomarkers with Cognitive Screening Test Performance in the Congolese Population in Kinshasa

**DOI:** 10.1101/2023.08.28.23294740

**Authors:** Megan Schwinne, Alvaro Alonso, Blaine R. Roberts, Sabrina Hickle, Inge MW Verberk, Emmanuel Epenge, Guy Gikelekele, Nathan Tsengele, Immaculee Kavugho, Samuel Mampunza, Kevin E. Yarasheski, Charlotte E. Teunissen, Anthony Stringer, Allan Levey, Jean Ikanga

## Abstract

**Background:** Alzheimer’s Disease (AD), the most common cause of dementia, poses a significant global burden. Diagnosis typically involves invasive and costly methods like neuroimaging or cerebrospinal fluid (CSF) biomarker testing of phosphorylated tau (p-tau) and amyloid-β_42/40_ (Aβ_42/40_). Such procedures are especially impractical in resource-constrained regions, such as the Democratic Republic of Congo (DRC). Blood-based biomarker testing may provide a more accessible screening opportunity.

**Objective:** This study aims to examine if AD-related blood-based biomarkers are associated with cognitive test performance in the Congolese population, where limited research has been conducted.

**Methods:** In this cross-sectional study of 81 Congolese individuals, cognitive assessments (Alzheimer’s Questionnaire (AQ) and Community Screening Interview for Dementia (CSID)) distinguished dementia cases from controls. Blood draws were taken to assess p-tau 181 and Aβ_42/40_ biomarkers. Relationships between the biomarkers and cognitive performance were analyzed using multiple linear regression models.

**Results:** Lower plasma Aβ_42/40_ was significantly associated with lower CSID scores and higher AQ scores, indicative of AD (p<0.001). These relationships were observed in healthy controls (CSID p=0.01, AQ p=0.03), but not in dementia cases. However, p-tau 181 did not exhibit significant associations with either measure. Factors such as age, sex, education, presence of APOE e4 allele, did not alter these relationships.

**Conclusion:** Understanding relationships between AD-related screening tests and blood-biomarkers is a step towards utilization of blood-based biomarker tests as a screening tool for AD, especially in resource-limited regions. Further research should be conducted to evaluate blood biomarker test efficacy in larger samples and other populations.

## INTRODUCTION

Dementia, one of the top five causes of death globally, is an umbrella term encompassing a group of characteristic symptoms, which include difficulties with memory, language, problem-solving, and other thinking skills.^1,2^ Alzheimer’s Disease (AD), the leading cause of dementia in individuals aged 65 or older, is a large global burden, with about 60 million people currently living with dementia^1^ and predicted to be over 150 million by 2050.^3,4^ Dementia is a clinical syndrome that does not require a diagnosis via biomarker measurements or PET-scans. However, AD is diagnosed based on clinical symptoms of dementia along with a distinct biomarker profile or PET-scan.^5^ AD is a slowly progressive neurodegenerative disease characterized by cerebral accumulation of amyloid plaques and neurofibrillary tangles.^4^ The changes in the brain from AD, such as the degeneration of nerve cells as well as the accumulation of the abnormal proteins, beta-amyloid and phosphorylated tau, are contributors to dementia.^2^ While the underlying cause of these pathological changes in AD is still unknown, the predominant risk factor is aging, in which the incidence is higher with increasing age with a late onset of 65 years or older. Genetic factors also play a major role, with 70% of AD cases relating to genetics.^4^ In fact, the e4 allele of the apolipoprotein E (APOE) gene is the strongest genetic risk factor for AD and the APOE e2 allele is the strongest genetic protective factor.^6^

As AD risk increases with age, it is important to focus on the African population, since they are aging at an unprecedented rate.^2,7^ This demographic transition is occurring faster in low and middle-income countries (LMIC) than it was in the previous century for high-income countries (HIC).^8^ Thus, the largest proportion of the predicted increase in AD will take place in LMIC, especially East Asia and Sub-Saharan Africa, where over 70% of individuals with dementia are expected to live in 2040.^7,8^

Common measures to screen for AD and related dementias include the Alzheimer’s Questionnaire (AQ) and the Community Screening Interview for Dementia (CSID). These measures combine a short cognitive screener with information from close contacts regarding daily functioning; the combination of the two types of tests yields better sensitivity and specificity for dementia diagnosis. ^9^ While the two screening tools have some different cognitive domains being tested, they both encompass semantic, executive, and memory knowledge.^10^ Since both assessments have different attributes that may be advantageous to different populations, it is important to utilize multiple screening tools and supplementary tactics to identify cases of AD.

Given that neurodegenerative diseases, such as AD, are difficult to diagnose clinically, characteristic biomarkers of AD, such as total tau (T-tau), phosphorylated tau (p-tau), amyloid- β_42_ (Aβ_42_), and amyloid-β_40_ (Aβ_40_), are important for research and early diagnosis.^11,12^ Increased levels of T-tau and p-tau with decreased Aβ_42_ in cerebrospinal fluid (CSF) is the biomarker pattern known as the “Alzheimer’s CSF Profile”, as they reflect key elements of AD pathophysiology.^13^ Tau protein normally binds to and stabilizes the neuronal microtubules but hyperphosphorylation disrupts the microtubules, impairs the plasma and axon flow, and leads to loss of neuronal connectivity.^13^ A lower Aβ_42_ reflects aggregation and deposition of protein in the brain. Aβ_40_ is the most abundant variant of Aβ in CSF, so the Aβ_42/40_ ratio is utilized to compensate for inter-individual differences in amyloid precursor protein (APP) expression and processing that can result in different but proportional concentrations of the CSF and plasma Aβ peptides. As such, the Aβ_42/40_ ratio is a better predictor of the presence of brain amyloid plaques than the plasma concentrations alone.^13,14,15^ Low CSF and plasma Aβ_42/40_ ratios and high tau concentrations are fluid biomarkers for AD pathology ^13^Although obtaining the biomarkers via CSF has been customary practice, obtaining the biomarkers from blood is more accessible than CSF and is preferable for both screening and sampling purposes.^13^ While there are several caveats making blood more challenging than CSF for brain biomarkers, such as dilution with other plasma proteins and degradation by proteases in the blood, novel developments in ultrasensitive immunoassays as well as mass spectrometry bring promising results for the use of blood biomarkers over CSF biomarkers.^13^

Substantial research has been implemented to demonstrate the use of cognitive tests as well as CSF biomarkers, such as Aβ_42/40_ and p-tau 181, to screen for AD; however, current research on the association between AD diagnosis and these biomarkers in blood has been predominantly limited to studies conducted in high-income countries. Very few studies have occurred in Sub-Saharan Africa, specifically in the DRC^7,8^. Given the invasiveness and expense of retrieving biomarkers via CSF, it is critical to determine other plausible screening methods to evaluate individuals for AD. Furthermore, with the increasing prevalence of AD in LMIC, in specific Africa, it is important to focus research on these populations, especially since the majority of AD-related research is conducted in populations of European ancestry and in high-income countries. This study aims to evaluate the association between AD-related plasma biomarkers with the cognitive tests, CSID and AQ, in a cohort from Sub-Saharan Africa.

## METHODS

### Study Design and Participants

From 2019 to 2022, a cross-sectional study using community-based recruitment was carried out in Kinshasa, the capital of the Democratic Republic of Congo (DRC). 1432 individuals were recruited from churches, clinics, hospitals, door-to-door, and older adult associations to then be screened. Eligibility criteria required that participants are 50 years or older, have a close contact to serve as a collateral informant, have no current or past history of neurodevelopmental, mental, psychiatric, or neurogenerative diagnosis other than dementia, able to give informed consent, fluent in French or Lingala, and have adequate sensory perceptual skills to be able to see and draw for cognitive tests. Cognitive test data was collected between 2019-2021 and blood specimens for biomarker analysis were collected between 2021-2022. Only some participants, less than those who had cognitive tests, were given the option to proceed with donating blood specimens. The enrollment flowchart is presented in Figure 1. This study was approved by the Ethics Committee and Institutional Review Boards of the University of Kinshasa and written informed consent was obtained from participants as well as financial compensation.

**Figure 1.**
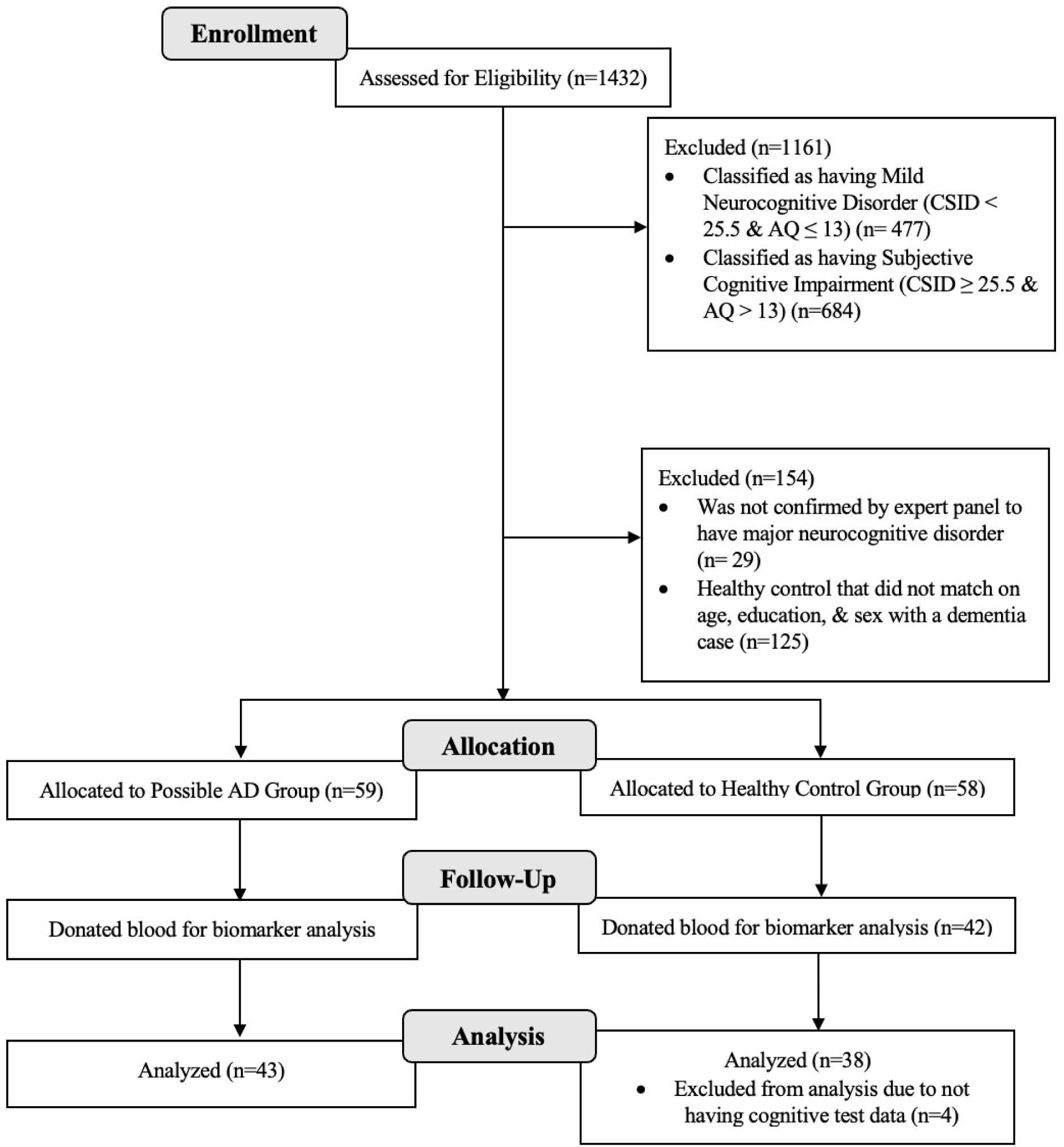
Flow Chart of Recruitment Status from those assessed for eligibility at enrollment (n=1432) to the individuals that were allocated to the dementia or control group and analyzed (n=81)

### Cognitive Measurements

Participants and their informants were administered the CSID as well as the AQ test to screen for dementia and to be further assigned into the dementia group or the healthy control group, which comprised of individuals with normal cognitive aging. The CSID, a 42-question screening measure, provides a score ranging from 0 to 55, with a lower score indicating worse cognition. As a widely accepted dementia screening tool to use cross-culturally, it serves to detect dementia in various populations with diverse educational, cultural, and linguistic identities.^7,9^ While this instrument is not the gold standard for diagnosis, it has been used in developing countries when higher quality screening instruments are not available. It evaluates the cognitive domains of language and expression, memory, learning, attention and calculation, praxis, orientation in space and time, and language comprehension. The AQ, an informant-only questionnaire, is 21-questions, with a score ranging from 0 to 26; a higher score indicates worse cognition. It is advantageous in providing questions that require yes or no answers in a weighted format, requiring no interpretation for individual components of the test.^10^ This assessment evaluates the memory, orientation, functional ability, visuospatial, and language domains. Participants were first classified using CSID scores as cognitively impaired (CSID score of < 25.5) or as cognitively unimpaired (CSID score of ≥ 25.5). Next, participants were classified within each category of cognition via AQ scores as cognitively impaired (AQ score of > 13) or as cognitively unimpaired (AQ score of ≤ 13). Given the two cognitive tests and classifications, 4 separate groups were created, which were major neurocognitive disorder (CSID < 25.5 and AQ > 13), mild neurocognitive disorder (CSID < 25.5 and AQ ≤ 13), subjective cognitive impairment (CSID ≥ 25.5 and AQ > 13), and normal cognition (CSID ≥ 25.5 and AQ ≤ 13), following DSM-IV terminology. Only the individuals with major neurocognitive disorder, which were considered to have dementia, and the individuals with normal cognition, which were considered healthy controls, were included for this analysis. Out of the 1432 initial participants, 271 met the above criteria for major neurocognitive disorder or normal cognition, in which 88 individuals were classified as having major neurocognitive disorder and 183 individuals were classified as healthy subjects with normal cognition. Following this classification, an expert panel of neuropsychologists, neurologists, and psychiatrists further evaluated the individuals through neurological and psychiatric evaluations, as well as assessing medical history. They then confirmed 55 individuals to have major neurocognitive disorder and then matched 59 healthy controls on age, education, and sex (Figure 1). Due to the participants not having PET-scans or CSF biomarker tests in this study, participants with major neurocognitive disorder will be characterized as having dementia and will not be identified as individuals with possible AD.^5^

### Descriptive Measurements

Participants were given self-report questionnaires and interviews to obtain demographic, socioeconomic, and medical history information. Individuals were categorized into age groups of 50-64, 65-74, 75-84, and 85+. Education levels were also categorized into levels of primary school (1-6 years), secondary school (7-12 years), some or completion of university (13-17 years), and beyond university (18+ years). Medical residents measured hypertension by using a manual sphygmomanometer and three measurements of systolic and diastolic blood pressure were collected. Having an average systolic blood pressure over 140 mmHg or a diastolic blood pressure over 90 mmHg was defined as hypertension.

### Biomarker Measurements

For the individuals that consented to blood donation for blood biomarker measurements, a phlebotomist drew the blood at the Medical Center of Kinshasa (CMK) blood laboratory by venipuncture into ethylenediaminetetraacetic acid (EDTA) tubes. Blood samples were centrifuged within 15 minutes and 5 ml plasma were aliquoted into 0.5 ml tubes. The samples were temporarily stored at -20 ° Celsius for less than a week and then at -80 ° C for longer-term storage at the CMK laboratory freezer. The samples were then shipped on dry ice to Emory University laboratory and analyzed by C_2_N Diagnostics (Aβ_42_ and Aβ_40_ peptides) and by Dr. Blaine Roberts’s lab at Emory University (p-tau 181). For the Aβ_42/40_ ratios, plasma samples were spiked with stable isotope labeled recombinant proteins. Plasma proteins were extracted using proprietary antibodies conjugated to magnetic beads, eluted from the beads, and then digested with a site-specific protease to form C-terminal peptides specific to Aβ_42_ and Aβ_40_ proteins. The peptides were separated using micro-flow liquid chromatography and electro-sprayed into the source of a high resolution orbitrap mass spectrometer. This procedure identifies the peptides of interest based on known amino acid sequence and mass to charge ratio. It then quantifies the ion signal intensity from the endogenous peptides by comparison to a calibration curve created with a stable isotope labeled internal standard peptide. The Aβ_40_ and Aβ_42_ concentrations were quantified by comparing the signal intensities for the endogenous peptides to those obtained from the stable isotope labeled proteins spiked into the sample. Aβ_42/40_ concentration ratios were calculated as plasma Aβ_42_ (pg/mL)/Aβ_40_ (pg/mL). To analyze p-tau 181 concentrations, EDTA plasma samples were prepared according to manufacturer’s instructions from the p-tau 181 kit v2 (Quanterix Billerica, Massachusetts, USA). Samples were run in a single path. Plasma was thawed at room temperature for 45 minutes and then centrifuged at 5000xg for 10 minutes. The plasma samples were then diluted four times on bench and measured on the Simoa HDX platform. Mean intraassay coefficients of variation (CV) were below 10%.

### Statistical Analysis

Of the study population, 81 individuals had both biomarker and cognitive data (43 dementia cases and 38 healthy controls, Figure 1). Preliminary analysis involved obtaining frequencies and means of sex, age, education level, and basic medical history, for the overall sample population and the two groups of differing neurological status separately. Chi-square test for proportions and two-sample t-tests were utilized to evaluate significant differences between healthy controls and dementia cases. Multiple linear regression models (Table 4, 5) were utilized to analyze associations between cognitive test scores outcome variable) and blood biomarkers (main independent variable). Neurological status was also a primary indicator variable in a model (Table 2) analyzing associations between either cognitive tests or blood biomarkers (outcome variables) with status. Analyses considered the overall CSID and AQ scores, as well as domain-specific scores (executive, semantic, and memory) separately. The biomarkers evaluated were plasma p-tau 181 and Aβ_42/40_ values. These models analyzed associations overall as well as stratified by neurological status (dementia or healthy control). Aβ_42/40_ was modeled in 0.01 increments, its standard deviation, to represent more meaningful findings in relation to associations with cognitive test scores. All models controlled for age, sex, and education, as these covariates may be possible confounders and bias the measures of association. The results were expressed as β-coefficients with corresponding 95% confidence intervals. Tests for potential interactions between biomarkers and covariates, including sex, age, education, and APOE status were conducted to understand if these variables significantly affected the relationship between biomarkers and cognitive tests. Tests for interaction involving the variables age and education were assessed on a continuous scale. The presence of the e4 allele in APOE genotypes, a known risk factor for AD, was assessed as a categorical variable, in which individuals were dichotomized as either having the e4 allele or not.^6^ All statistical tests were two-sided, and p-values < 0.05 were considered to be statistically significant. All analyses were conducted using SAS version 9.4 statistical software.

## RESULTS

### Descriptive and Clinical Characteristics of the Sample Population

Baseline characteristics of the 81 individuals, including demographics and medical history, were reported in Table 1. The sample population consisted of 43 dementia cases and 38 healthy controls with a mean age of 73 years (ranging from 50-88 years old). Sex, body mass index, age groups, and education levels were similar between the dementia and control groups, confirming matching was performed appropriately. Regarding medical history, a large proportion of the participants (53%) had hypertension, with more dementia cases having prevalent hypertension compared to the controls (60% and 45% respectively). Additionally, more of the dementia cases (28%) reported alcohol abuse compared to the control group (11%). The remaining relevant medical history and mental conditions, such as high cholesterol, poor nutrition, anxiety, and depression were minimally reported among the sample. The presence of at least one APOE e4 allele, and the specific APOE genotypes, significantly differed between healthy controls and dementia cases (p=.004). Overall, the prevalence of e4 allele was higher in dementia cases than healthy controls, with e3/e4 being the more common genotype in this group.

**Table 1.** Descriptive Characteristics of the Sample Population, Stratified by Neurological Status.

| Variable,<br>n (%) | Overall<br>(n = 81) | Healthy Controls<br>(n = 38) | Dementia Cases<br>(n = 43) | p-value |
| --- | --- | --- | --- | --- |
| **Demographics** |  |  |  |  |
| Male | 35 (43%) | 16 (42%) | 19 (44%) | 0.85 |
| Body Mass index, kg/m<sup>2</sup> * | 24.7 (4.3) | 24.6 (4.1) | 24.8 (4.4) | 0.86 |
| Obesity Status |  |  |  | 0.09 |
| Underweight (BMI < 18.5) | 2 (2%) | 2 (5%) | 0 (0%) |  |
| Healthy Weight (BMI ≥ 18.5, <25) | 45 (56%) | 18 (47%) | 27 (63%) |  |
| Overweight (BMI ≥ 25, <30) | 22 (27%) | 14 (37%) | 8 (19%) |  |
| Obese (BMI ≥ 30) | 12 (15%) | 4 (11%) | 8 (19%) |  |
| Age, years * | 73.0 (8.0) | 71.7 (7.9) | 74.0 (8.1) | 0.19 |
| Age Groups, years |  |  |  | 0.19 |
| 50-64 | 11 (14%) | 6 (16%) | 5 (12%) |  |
| 65-74 | 29 (36%) | 16 (42%) | 13 (30%) |  |
| 75-84 | 37 (46%) | 16 (42%) | 21 (49%) |  |
| 85+ | 4 (5%) | 0 (0%) | 4 (9%) |  |
| Years of Education * | 8.2 (5.4) | 9.2 (5.3) | 7.3 (5.5) | 0.12 |
| Education Level |  |  |  | 0.47 |
| Primary School (1-6 years) | 8 (10%) | 2 (5%) | 6 (14%) |  |
| Secondary School (7-12 years) | 28 (35%) | 12 (32%) | 16 (37%) |  |
| Some/Completed University (13-17 years) | 27 (33%) | 14 (37%) | 13 (30%) |  |
| Beyond University (18+ years) | 18 (22%) | 10 (26%) | 8 (19%) |  |
| **APOE e4** |  |  |  |  |
| Presence of $\geq 1$ e4 Allele † | 40 (50%) | 12 (32%) | 28 (65%) | 0.0036 |
| e2/e4 Genotype | 2 (3%) | 2 (5%) | 0 (0%) |  |
| e3/e4 Genotype | 30 (38%) | 9 (24%) | 21 (49%) |  |
| e4/e4 Genotype | 8 (10%) | 1 (3%) | 7 (16%) |  |
| **Medical History** |  |  |  |  |
| Hypertension † | 43 (54%) | 17 (46%) | 26 (60%) | 0.19 |
| High Cholesterol† | 2 (3%) | 1 (3%) | 1 (2%) | 0.94 |
| Poor Nutrition | 1 (1%) | 0 (0%) | 1 (2%) | 0.34 |
| Stroke† | 2 (3%) | 0 (0%) | 2 (5%) | 0.17 |
| Tobacco Abuse | 9 (11%) | 5 (13%) | 4 (9%) | 0.58 |
| Alcohol Abuse | 16 (20%) | 4 (11%) | 12 (28%) | 0.0499 |
| Anxiety | 5 (6%) | 2 (5%) | 3 (7%) | 0.75 |
| Depression | 7 (9%) | 2 (5%) | 5 (12%) | 0.29 |

\*This variable is reported as mean (SD)
†These values may not sum to the total due to missing data

### Descriptive Characteristics of Cognitive Tests

Upon comparison of CSID and AQ cognitive test scores between dementia cases and healthy controls (Table 2), all overall scores as well as the semantic, executive, and memory domain scores were significantly different between the two groups (p < 0.01). Given that the higher the CSID score, the better the cognition, on average, the healthy controls scored higher in all CSID domains compared to the dementia cases. The CSID memory domain had the largest difference between groups and the CSID semantic domain scores were the least impacted. Furthermore, the healthy control group scored lower on the AQ cognitive test compared to the dementia cases. Again, the AQ memory domain yielded the largest difference and the AQ semantic domain scores yielded the smallest difference between groups.

**Table 2.** Descriptive Data of Cognitive Tests and Biomarkers, stratified by Neurological Status.

| Variable,<br>Mean (SD) | Controls<br>(n = 38) | Dementia<br>(n = 43) | Overall<br>(n = 81) | $\beta_1$ (95% CI) * | p-value |
| --- | --- | --- | --- | --- | --- |
| **Cognitive Tests** |  |  |  |  |  |
| **CSID** |  |  |  |  |  |
| Overall Score | 31.1 (4.2) | 19.7 (5.6) | 25.0 (7.6) | -11.0 (-13.2, -8.8) | <.001 |
| CSID Semantic Domain Score | 9.9 (0.3) | 8.7 (1.8) | 9.3 (1.5) | -1.2 (-1.8, -0.6) | 0.003 |
| CSID Executive Domain Score | 14.9 (2.0) | 10.0 (3.8) | 12.3 (3.9) | -4.6 (-6.0, -3.3) | <.001 |
| CSID Memory Domain Score | 20.7 (4.3) | 10.6 (5.1) | 15.3 (6.9) | -9.9 (-12.0, -7.8) | <.001 |
| **AQ Test** |  |  |  |  |  |
| Overall Score | 4.3 (5.4) | 19.1 (3.9) | 12.1 (8.8) | 14.7 (12.6, 16.9) | <.001 |
| AQ Semantic Domain Score | 0.2 (1.0) | 2.3 (1.7) | 1.3 (1.7) | 2.0 (1.4, 2.7) | <.001 |
| AQ Executive Domain Score | 1.4 (1.7) | 6.3 (1.8) | 4.0 (3.0) | 4.8 (4.0, 5.6) | <.001 |
| AQ Memory Domain Score | 2.7 (3.0) | 10.4 (2.7) | 6.8 (4.8) | 7.8 (6.5, 9.1) | <.001 |
| **Blood Biomarkers** |  |  |  |  |  |
| A$\beta_{42/40}$ | 0.106 (0.009) | 0.099 (0.008) | 0.102 (0.009) | -0.006 (-0.009, -0.002) | 0.002 |
| A$\beta_{42}$, pg/ml | 51.0 (10.8) | 47.8 (11.8) | 49.3 (11.4) | -3.8 (-8.9, 1.4) | 0.152 |
| A$\beta_{40}$, pg/ml | 486.0 (105.1) | 483.4 (117.0) | 484.6 (111.0) | -11.7 (-62.5, 39.0) | 0.647 |
| p-tau 181, ng/ml | 1.5 (1.4) | 1.6 (1.4) | 1.6 (1.4) | 0.02 (-0.6, 0.7) | 0.939 |

\*Results from linear regression models are adjusted for age, sex, and education.

### Descriptive Characteristics of Blood Biomarkers

Average, Aβ_40_, Aβ_42_, Aβ_42/40_ ratio, and p-tau 181 measures are also presented in Table 2. Aβ_42/40_ was significantly higher in the control group compared to the dementia group (p=0.002). Aβ_40_ and Aβ_42_ were not significantly different between groups, but the ratio is most clinically relevant as a biomarker for AD pathology. While Aβ_42/40_ was significantly different, analysis of p-tau 181 yielded essentially identical concentrations between the two groups of differing neurological statuses (p=0.94).

**Table 3.** Assessing Interaction between Biomarkers and Covariates.

| Test | Biomarker | p-value for Interaction |  |  |  |
| --- | --- | --- | --- | --- | --- |
|  |  | Age | Sex | Education | APOE e4 Allele |
| CSID Total Score | p-tau 181, ng/ml | 0.57 | 1.00 | 0.17 | 0.17 |
| | A$\beta_{42/40}$ † | 0.76 | 0.43 | 0.41 | 0.27 |
| Overall AQ Score | p-tau 181, ng/ml | 0.80 | 0.21 | 0.30 | 0.18 |
| | A$\beta_{42/40}$ † | 0.45 | 0.24 | 0.33 | 0.07 |

† A $\beta_{42/40}$ is modeled in 0.01 increments.

### Association between Cognitive Tests and Biomarkers

Upon exploration of potential associations between blood biomarkers and cognitive test scores among the whole study population, Aβ_42/40_ was strongly associated with both CSID and AQ overall scores (p<0.001), while p-tau 181 was not. CSID overall scores and Aβ_42/40_ demonstrated a positive association while AQ overall scores and Aβ_42/40_ demonstrated a negative association. For every 0.01 increase in Aβ_42/40_, on average the CSID overall score was 3.77 points higher, after adjusting for age, sex, and education. Furthermore, for every 0.01 increase in Aβ_42/40_, on average the AQ overall score was 4.58 points lower (a cognitively better score) after adjustment. Aβ_42/40_ was only significantly associated with the CSID test (p=0.01) and the AQ test (p=0.03) among the healthy controls and not among the individuals with dementia (Table 4). Potential interaction between biomarkers with covariates, including sex, age, education, and APOE status was assessed. The test for interactions between these variables all resulted in p-values > 0.05, meaning interaction was not present and these variables did not modify the relationship between cognitive tests and biomarkers.

**Table 4.**
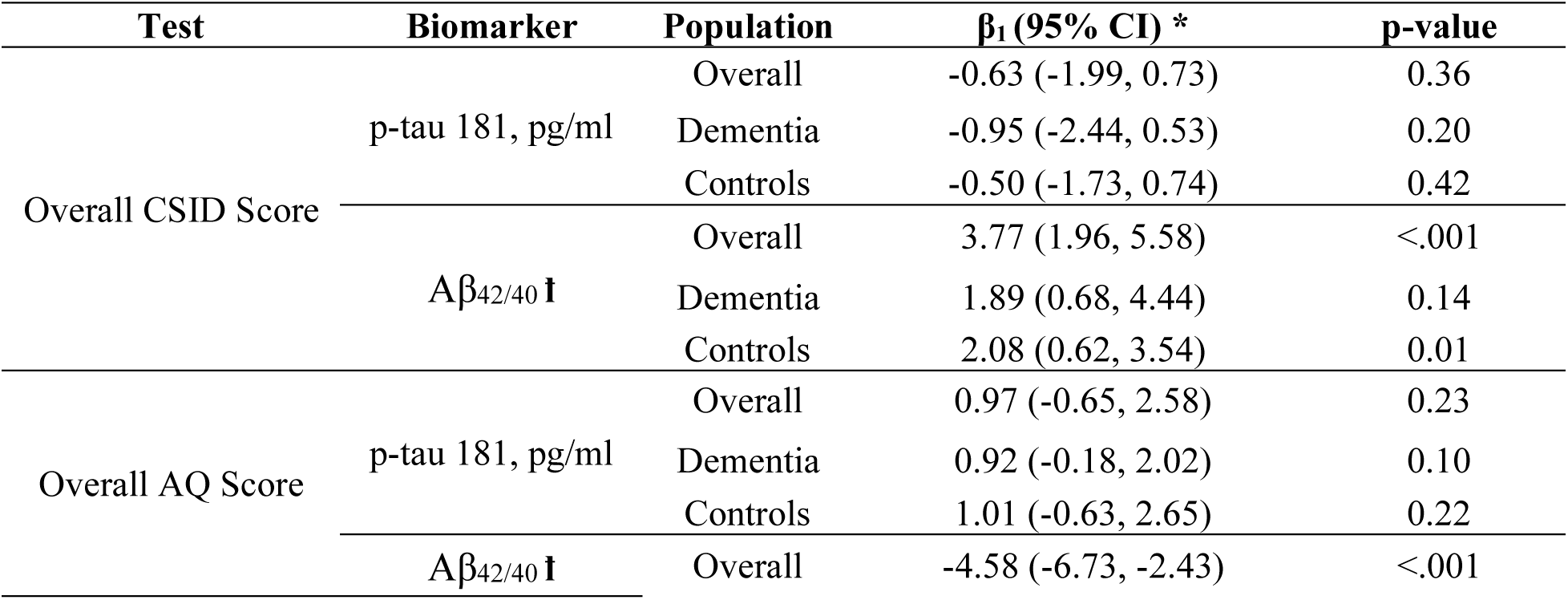
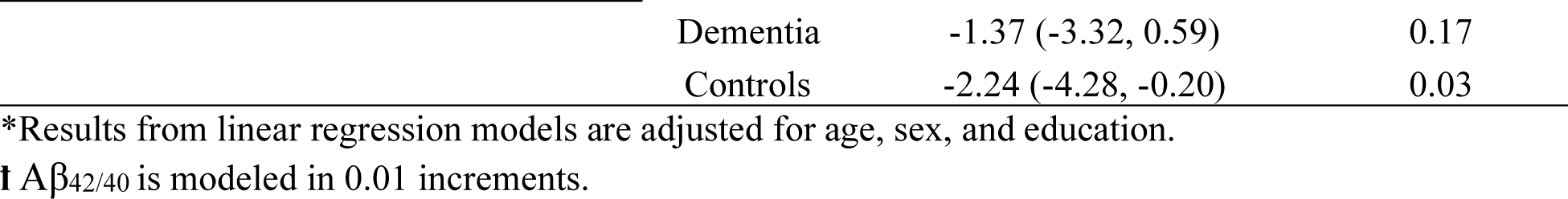
Association Between Cognitive Tests and Biomarkers, Overall and Stratified by Neurological Status.

Cognitive tests were also stratified into their 3 domains, semantic, executive, and memory function (Table 5). Only the CSID semantic domain was associated with p-tau 181 (p=0.03), while all other domains were not significantly associated with this biomarker. However, all cognitive test domains were associated with Aβ_42/40_ (p≤0.001) except for the CSID semantic domain. For both the CSID and AQ tests, the memory domain had the strongest difference by 0.01 Aβ_42/40_ increments (3.47 and -2.4 respectively), while the semantic domain had the smallest rate of change (0.38 and -0.78 respectively).

**Table 5.** Association Between Cognitive Test Domains and Biomarkers.

| Test | p-tau 181, ng/ml | | A$\beta_{42/40}$ † | |
| --- | --- | --- | --- | --- |
| | $\beta_1$ (95% CI) * | p-value | $\beta_1$ (95% CI) * | p-value |
| CSID |  |  |  |  |
| Overall Score | -0.63 (-2.0, 0.7) | 0.36 | 3.77 (2.0, 5.6) | <.001 |
| Semantic Domain Score | -0.30 (-0.6, -0.02) | 0.03 | 0.38 (-0.01, 0.8) | 0.06 |
| Executive Domain Score | -0.51 (-1.2, 0.2) | 0.15 | 1.78 (0.9, 2.7) | <.001 |
| Memory Domain Score | -0.20 (-1.5, 1.1) | 0.75 | 3.47 (1.8, 5.1) | <.001 |
| AQ |  |  |  |  |
| Overall Score | 0.97 (-0.6, 2.6) | 0.23 | -4.58 (-6.7, -2.4) | <.001 |
| Semantic Domain Score | 0.20 (-0.1, 0.5) | 0.23 | -0.78 (-1.2, -0.3) | <.001 |
| Executive Domain Score | 0.29 (-0.3, 0.8) | 0.29 | -1.27 (-2.0, -0.5) | 0.001 |
| Memory Domain Score | 0.39 (-0.5, 1.3) | 0.39 | -2.4 (-3.7, -1.2) | <.001 |

\*Results from linear regression models are adjusted for age, sex, and education.
† A $\beta_{42/40}$ is modeled in 0.01 increments.

## DISCUSSION

### Major Findings

In a community-based sample from the DRC, we found associations of blood Aβ_42/40_ with CSID and AQ scores, with lower Aβ_42/40_ correlating with a lower CSID score and higher AQ score, which is characteristic of AD and other dementias. These relationships were present for healthy controls but not participants with dementia. However, circulating p-tau 181 was not associated with either cognitive test. Age, sex, education, and presence of the APOE e4 allele, which are known risk factor for AD, did not significantly modify these associations.

Previous research suggests that the use of biomarkers alongside neurocognitive tests is the future of clinical practice, as they aid in early identification of AD and potential for prevention of dementia manifestation or progression.^3^ The literature has repeatedly shown that reduced Aβ_42/40_ and increased p-tau is characteristic of AD, and these relationships have commonly been seen in CSF and more recently studied in blood.^13,15^ Plasma Aβ_42/40_ had similar results with CSF tests, however, p-tau did not align as clearly and showed a weaker relationship.^16^ This supports these present findings since Aβ_42/40_ was associated with tests displaying cognitive impairment in multiple domains, but p-tau was not. With that said, a recent study investigating associations between these plasma biomarkers and their relationship with AD-associated neuroimaging results show that both biomarkers are significantly associated with AD.^17^

Memory significantly differs among those with dementia most likely due to pathophysiological changes, such as the accumulation of amyloid-beta and the development of hyperphosphorylated tau protein tangles.^3^ This then leads to secretion of neurotoxins and inflammatory factors, resulting in neuronal death in specific brain areas and causes memory impairment.

Aβ_42/40_’s apparent association compared to p-tau 181’s insignificant association with cognitive status may be due to the differences in pathophysiological processes between the biomarkers. Aβ_42/40_ is an early, pre-symptomatic index for increased neurotoxicity, amyloidogenicity, as well as disease severity, whereas p-tau 181 may be a more delayed, symptomatic index for tau hyperphosphorylation, neurofibrillary tangles formation, and degenerative axonal loss in the brain.^17^ These processes may manifest differently in blood samples compared to CSF samples, differ between individuals, or differ by the tau phosphorylation site (p-tau181, 217, 231, etc.) measured, since the sites may have slightly different temporal patterns over the disease course. Moreover, it is possible that the significant associations of Aβ_42/40_ but not p-tau 181 is due to higher performance of the mass spectrometry Aβ_42/40_ assay compared to the p-tau 181 assay. It is interesting to note that an earlier study among predominantly asymptomatic African Americans showed that plasma Aβ_42/40_ measures were more highly correlated with brain amyloid status (PET and CSF) than the p-tau 181 biomarker.^18^ The findings from the current DRC cohort, support this observation, and suggest the possibility that plasma tau biomarkers may need to be interpreted differently in people of African descent vs. non-Hispanic Caucasians. It is also possible that the lack of association between p-tau 181 and cognitive status in this study was due to the small sample size and may not represent a true finding.

While blood is cheaper, less invasive, and more accessible to sample from individuals compared to CSF, there are inevitable caveats to blood biomarker analysis.^19^ It is more difficult to reliably measure blood biomarkers that are related to cognitive disorders because the biomarkers are present at lower concentrations in the blood compared to CSF, which is closer to the brain and allows for a free exchange of molecules.^13,16,20^ Only a fraction of brain proteins enters the blood stream and biomarker dilution from peripheral sources may occur. Brain proteins released in the blood may be degraded by proteases or metabolized in the liver, leading to potential for varying measurements that may not necessarily be representative of brain or CSF biomarker levels, and thus, cognitive impairment. Lastly, the low levels of brain proteins entering the blood are mixed in a matrix containing high levels of unrelated plasma proteins that need to be cleared from the plasma during sample preparation and may skew results.^13^ Nevertheless, advancements in technology have aided in showing the feasibility of measuring blood-based biomarkers more accurately and with clinically meaningful results.

### Strengths and Limitations

This study had numerous strengths through the study design, setting, and statistical analyses methods. First, Sub-Saharan Africa, specifically the DRC, is an understudied area for dementia and AD, therefore this research strengthens and adds to knowledge of AD and its associated biomarkers for this population. The research staff were also familiar with the area and the population of interest so there was enhanced partnership and no language barrier. Third, the sample was relatively healthy and varied in age, sex, and education levels. Additionally, multiple cognitive tests (CSID and AQ) as well as an expert panel were utilized to establish and confirm the participants cognitive ability to prevent misclassification. Additionally, dementia cases and healthy controls were matched on age, sex, and education as part of the study design, but these covariates were also controlled for in analysis, decreasing the possibility of confounding. Lastly, statistical tests prevented further potential bias by confirming there were no outliers or key variables causing interaction.

As with all research studies, there are inevitably limitations. While Sub-Saharan Africa, specifically the DRC is an understudied area, the population within the region lacks variability in race and ethnicity, so there is lack of generalizability to other populations. Additionally, given the location, it is a complicated setting with less access to advanced technology. Variables that may have been risk factors or confounders, such as physical activity and family history were not considered and the sample size of 81 is rather low, decreasing the statistical power. Lastly, there was a discrepancy in the number of questions in this study’s CSID test compared to the most common CSID test (36 vs. 42 questions), which decreases comparability.

### Conclusions

Understanding the AD-specific blood biomarkers Aβ_42/40_ and p-tau 181 relationships with neurocognitive tests related to AD is a promising next step in the implementation of blood-based biomarkers in order to overcome access and cost barriers, especially in LMIC. Instruments for quantifying blood biomarkers are becoming more sensitive and implementation is increasing. While blood biomarkers are not equivalent to an AD diagnosis yet, they can be utilized as a screening tool before resorting to PET-scan neuroimaging or CSF biomarker analysis. Future studies are needed in which AD-related blood and CSF biomarkers are tested longitudinally from the same individuals for better comparison and further validation. Furthermore, larger studies with greater sample sizes, additional biomarkers, and diversity in races and ethnicities should be employed to increase global generalizability.

## ACKNOWLEDGEMENTS

The authors acknowledged the expertise from the Centre Médical de Kinshasa (CMK), who have assisted our study with blood specimen collection, as well as C2N Diagnostics Laboratory for support in plasma biomarker analysis. Additionally, the authors thank Emory Rollins School of Public Health for guidance with data processing and analysis. Furthermore, this research is Supported by the National Center for Advancing Translational Sciences of the National Institutes of Health under Award # UL1TR002378. The content is solely the responsibility of the authors and does not necessarily represent the official views of the National Institutes of Health.

## FUNDING

This study was supported by grants AARG-19-61778701 and P30AG066511-02S1 from the National Institute on Aging. Dr. Alonso was supported by NIH/NHLBI grant K24HL148521 and NIH/NIA grant P30AG066511. C_2_N Diagnostics analyses were supported by the NIH (grant No. R44 AG059489), BrightFocus (grant No. CA2016636), The Gerald and Henrietta Rauenhorst Foundation, and the Alzheimer’s Drug Discovery Foundation (grant No. GC-201711-2013978).

## CONFLICT OF INTEREST

KEY is employed by and receives equity compensation from C_2_N Diagnostics, LLC. All other authors have no conflict of interest to report.

## DATA AVAILABILITY

The data supporting the findings of this study are available on request from the corresponding author. The data are not publicly available due to privacy or ethical reasons.

## Notes

### Competing Interest Statement

KEY is employed by and receives equity compensation from C2N Diagnostics, LLC. All other authors have no conflict of interest to report.

### Funding Statement

This study was funded by grants AARG-19-61778701 and P30AG066511-02S1 from the National Institute on Aging. Dr. Alonso was supported by NIH/NHLBI grant K24HL148521 and NIH/NIA grant P30AG066511. Dr. Daniel Drane and the neuroimaging analysis efforts were funded by the NIH/NINDS through grant R01 NS088748. C2N Diagnostics analyses were supported by the NIH (grant No. R44 AG059489), BrightFocus (grant No. CA2016636), The Gerald and Henrietta Rauenhorst Foundation, and Alzheimer's Drug Discovery Foundation (grant No. GC-201711-2013978).

### Author Declarations

Ethics Committee and Institutional Review Boards of the University of Kinshasa gave ethical approval for this work.

### Summary of Updates

Minor tweaks to Table 1, Figure 1, and text.

